# The impact of early anti-SARS-CoV-2 antibody production on the length of hospitalization stay among COVID-19 patients

**DOI:** 10.1101/2023.03.02.23286693

**Authors:** Dalziza Victalina de Almeida, Priscila Alves Cezar, Thais Freitas Barreto Fernandes, Marcos Gustavo Araujo Schwarz, Leila Mendonça-Lima, Carmem Beatriz Wagner Giacoia-Gripp, Fernanda Heloise Côrtes, Monick Lindenmeyer Guimarães, Jose Henrique Pilotto, Nathalia Beatriz Ramos de Sá, Andressa da Silva Cazote, Larissa Rodrigues Gomes, Marcel de Souza Borges Quintana, Marcelo Ribeiro-Alves, Lara Coelho, Kim Mattos Geraldo, Maria Pia Diniz Ribeiro, Sandra Wagner Cardoso, Beatriz Gilda Jegerhom Grinsztejn, Valdiléa Gonçalves Veloso dos Santos, Mariza Gonçalves Morgado

**Author notes:** Correspondence: Dalziza Victalina de Almeida - Address: Laboratory of AIDS & Molecular Immunology, Oswaldo Cruz Institute, FIOCRUZ. Av. Brazil 4365, Leonidas Deane Building, Room 401, Rio de Janeiro, Brazil. 21040-360 - -.

## Abstract

COVID-19 has challenged the scientific community in the search for biological markers and information that can contribute to the early management of the severe disease. Given the global scale of COVID-19, including reports of reinfection even in the presence of effective vaccines, we have not yet been able to eradicate the disease. This factor implies the emergence of new waves and an increasing number of hospitalizations. This study aimed to characterize the neutralizing antibody (Nab) geometric mean titers (GMTs) in hospitalized patients with COVID-19 and to evaluate the association with length of stay, comorbidities, and patient outcome. Among the 103 participants, 84 (81.5%) had some previous condition associated with worsening health, and 31 (30%) died. We found that neutralization potency varied greatly across individuals and was significantly higher in patients discharged before 14 days than in patients who stayed longer in the hospital. During the study period, 15 people living with HIV (PLWH) were hospitalized, and no significant difference in clinical characteristics or anti-SARS-CoV-2 Nabs was observed. However, PLWH with severe COVID-19 were younger (41.7, IQR=17.5) than other hospitalized COVID-19 patients (59.3, IQR=22, *P* <0.01). A high anti-HIV-1 antibody GMT of 583.9 (95% CI: 344-990) was detected, demonstrating maintenance of anti-HIV-1 Nab production among PLWH coinfected with SARS-CoV-2. Therefore, these results indicate that neutralizing antibodies are not the only immunological response capable of controlling disease progression. Nevertheless, these data highlight the importance of more Nab screening studies to predict shorter hospital stays.

## 1. Introduction

The major portal of SARS-CoV-2 entry into host cells is through the respiratory tract, with the virus binding to the angiotensin-converting enzyme 2 (ACE-2) receptor on alveolar type 2 (AT2) cells in the epithelium. When attacked by the virus, AT2 cells produce inflammatory mediators, stimulating proinflammatory chemokine production and initiating immunoglobulin production (1). Generally, immunoglobulin production occurs 4 days after the onset of symptoms, enabling the diagnosis of COVID-19 through specific SARS-CoV-2 antibodies (2). The detection of neutralizing antibodies (Nabs) against SARS-CoV-2 helps to evaluate the status of the immune response of asymptomatic/symptomatic COVID-19 patients and severe COVID-19, in addition to indicating protection correlates (3).

Recent studies have demonstrated that the magnitude of Nab responses appears to correlate with viral load, with higher responses reported among patients with more severe disease and older adults (4, 5). Several comorbidities, including respiratory disease, hypertension, diabetes, kidney disease, and HIV-1 infection, have been identified as risk factors associated with severe COVID-19 manifestation (6). Nonetheless, few studies describing the anti-SARS-CoV-2 Nab response in HIV/COVID-19 coinfections have been performed thus far (7). The identification and characterization of Nabs can help prevent new infections in exposed individuals, and passive immunization can be used as a treatment against disease progression to severe forms (8, 9).

The main objective of this study was to provide preliminary data on the Nab immune response to SARS-CoV-2 in a group of hospitalized subjects with COVID-19 in Rio de Janeiro, Brazil, including those with COVID-19 associated with HIV-1 infection. For this, we calculated the seroconversion rate of SARS-CoV-2 Nabs using the pseudovirus neutralization assay (PNA) (10) for a cohort of 103 hospitalized COVID-19 patients with or without HIV-1-associated infection and evaluated its association with clinical features of COVID-19.

## 2. Materials and Methods

### Human samples

Plasma samples analyzed in this study were obtained from a clinical COVID-19 follow-up study that included moderate/severe hospitalized patients at the COVID-19 Pandemic Hospital Center INI/FIOCRUZ, Rio de Janeiro, Brazil, between June 2020 and May 2021 (RECOVER-SUS study-NCT04807699). A total of 103 hospitalized participants who tested positive for SARS-CoV-2 by nasopharyngeal sampling using RT–PCR were selected. Plasma samples were collected on the first day of hospitalization (D1), after 14 days (D14), and between seven and eleven months (D300) after hospital discharge. None of the patients were vaccinated for COVID-19 during the blood collection period. The local ethics committee approved this study, and all participants signed an informed consent form; all the samples were deidentified.

### Cells

HEK293T-ACE2 and HEK293T/17 cells were cultured in DMEM with 10% fetal bovine serum (FBS), 2 mM of L-glutamine, and 200 μg/mL of hygromycin B (Thermo Fisher Scientific, US) at 37 °C with 5% CO_2_. HEK293T-ACE2 cells were a donation from BEI resources (BEI catalog number NR-52511), and the TZM-bl cell line was obtained through the NIH AIDS Research and Reference Reagent Program.

### Pseudovirus production and titration

All transfections were performed in T75 culture flasks, and the pseudovirus was produced in HEK293T/17 cells cultured 24 hours before reaching 50%–70% confluence on the day of the experiment. Viral particles were produced using the Luciferase-IRES-ZsGreen backbone obtained from the BEI resource, which was established by Crawford et al., 2020 (10). Briefly, 1 μg of HDM-IDTS pike-fixK (BEI catalog number NR-52514), 6 μg of pHAGE-CMV-Luc2-IRES-ZsGreen-W (BEI catalog number NR-52516), 1.4 μg of HDM-Hgpm2 (BEI catalog number NR-52517), 1.4 μg of HDM-tat1b (NR-52518) and 1.4 μg of pRC-CMV-Rev1b (NR-52519) were used and cotransfected with Lipofectamine 3000 (Thermo Fisher Scientific, US) and Opti-MEM I Reduced Serum Medium (Gibco) was used, as well. For the positive control, VSV G-pseudotyped lentiviral particles with the ZsGreen backbone were produced using the plasmid pHEF expressing vesicular stomatitis virus (VSV-G), obtained from the NIH-HIV Reagent Program (catalog number RP-4693). Pseudovirus supernatants were collected approximately 72 h post-transfection, and the FBS concentration in the virus-containing culture medium was adjusted to 20% (i.e., for each 1 ml of virus harvested, 0.125 ml of FBS was added), filtered through a 0.45-μm filter, and stored at -80 °C.

For median tissue culture infectious dose (TCID_50_) assay measurements, 293T-ACE-2 cells (4 × 10_4_cells in 100 µL of growth medium/well) were added to each well, followed by 20 mg/ml of DEAE-dextran with 11 pseudovirus dilutions, added after 24 h. Serial 5-fold dilutions of pseudovirus were quadruplicated in 96-well culture plates in a volume of 100 µL of growth medium/well. After 48 h of incubation (37 °C in 5% CO_2_), 100 µL of culture medium was removed from each well, and 100 µL of Britelite Plus Reagent (PerkinElmer) was added to the cells. After 2 min of incubation at room temperature to allow cell lysis, 150 µL of the cell lysate was transferred to black, solid 96-well plates for luminescence measurements using a GloMax® Navigator Microplate Luminometer (Promega, US). Wells with pseudovirus SARS-CoV-2-containing supernatant that was not toxic to the cells based on light microscopy inspection, and that produced at least relative luminescence units (RLU) 10 times greater than the background were scored as positive. The TCID_50_was calculated using the Reed–Muench method and using the macro described on the TCID_50_; the cutoff macro is available on the Los Alamos HIV Immunology Database (https://www.hiv.lanl.gov/content/nab-reference-strains/html/home.htm).

### Pseudovirus Neutralization Assay

Neutralization activity was expressed as Nab titers, defined as the interpolated plasma dilution that produced a 50% reduction in virus infectivity or a 50% (ID_50_) and 90% inhibitory dose (ID_90_), observed as reductions in Luciferase reporter gene expression after a single round of virus infection of 293T-ACE-2 cells, given that some components essential for viral replication or persistent infection were deleted from the genome. For each sample, residual infection was estimated across eight serum dilutions ranging from 1:20 to 1:4,3740; 3-fold serial dilutions of samples were performed in duplicate (96-well flat bottom plate) in DMEM growth medium (100 µL/well). An amount of virus to achieve a TCID_50_of 100 000 RLU equivalents was added to each well in a volume of 50 µL. The plates were then incubated for 90 min at 37 °C in a total volume of 150 µL of growth medium. 293T-ACE-2 cells (4 × 10^4^cells in 100 µL of growth medium) were added to each well, and after 24 h, 20 mg/ml of DEAE-dextran was added. The normalized % inhibition was calculated using uninfected cells (100% inhibition), and as a negative control, we used prepandemic plasma (0% inhibition) as a reference. For specificity, we used anti-SARS coronavirus recombinant human IgG1 (BEI catalog number: NR-52392). After 48 h of incubation, 150 µL of culture medium was removed from each well, and 100 µL of Britelite reagent (PerkinElmer, US) was added to the cells. After 2 min of incubation at room temperature to allow cell lysis, 150 µL of the cell lysate was transferred to black, solid 96-well plates for luminescence measurements using a GloMax® Navigator Microplate Luminometer (Promega, US). The assays were evaluated as having passed when the following parameters were met: the mean RLU of the virus control was 10 × higher than the background of the cell control; and the standard deviation of RLU in the virus control well was 30%. All data were analyzed using Excel Macros Nab analysis provided by the Global HIV Vaccine Enterprise, available on the Los Alamos HIV Immunology Database (https://www.hiv.lanl.gov/content/nab-reference-strains/html/home.htm).

### Statistical analysis

Nonparametric Mann–Whitney U tests were used to compare continuous numerical baseline demographic and clinical variables, whereas Fisher’s exact tests were used for categorical variables. Multiple linear fixed effects models were used to evaluate patient differences related to clinical outcome (discharge or death) and HIV-1 serology. The model’s fixed systematic component was adjusted by confounding variables (age, sex, self-declared race, number of comorbidities, and days since first symptoms of COVID-19 at hospital admission). The mean and 95% confidence intervals (95% CIs) were used, and the results were also presented graphically for the estimated mean marginal effects, where all other variables included in the multiple linear fixed models remained in equal proportions or their average values, and contrasts were constructed from these estimated mean marginal effects. For each parameter (Figure 1), a nonparametric ANOVA (Sidak’s multiple comparisons test) was performed. Tukey’s honest significant difference (HSD) method was used to correct *p* values by the number of comparisons whenever applicable (Figure 2). R software (https://www.r-project.org/about.html) version 4.1.1 packages ‘lme4’, ‘emmeans’, and their dependencies were used to perform the statistical analyses. For each parameter, a nonparametric ANOVA was performed, and a paired t test was performed to compare Nab titers (Figures 3 and 4). To evaluate the continuous ID_50_and ID_90_outcomes, we used a simple linear regression model assuming normal distribution for these outcomes on the log scale. Statistical significance was declared at a *p value* ≤0.05. Analysis was performed with statistical computing software GraphPad Prism 9.

**Figure 1:**
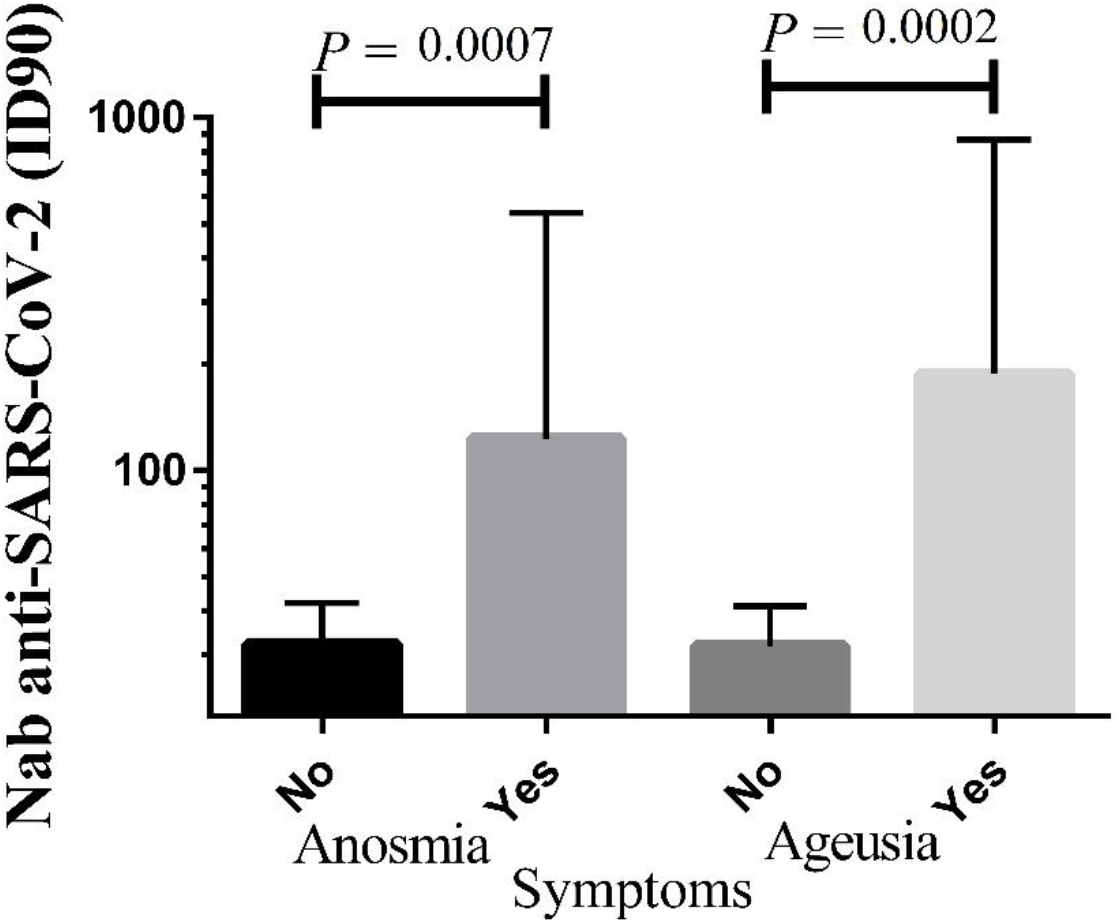
Clinical symptoms of anomia and ageusia of SARS-CoV-2 infection are influenced by neutralizing antibody titers. The COVID-19 patients (n = 103) were divided into groups according to the presence or absence of these symptoms and analyzed for their Nab titers with 90% inhibition (ID_90_). The bar line represents the geometric mean with a 95% confidence interval. For each parameter, a nonparametric ANOVA (Sidak’s multiple comparisons test) was performed; statistical significance is indicated with the following notation: *P*<0.001.

**Figure 2:**
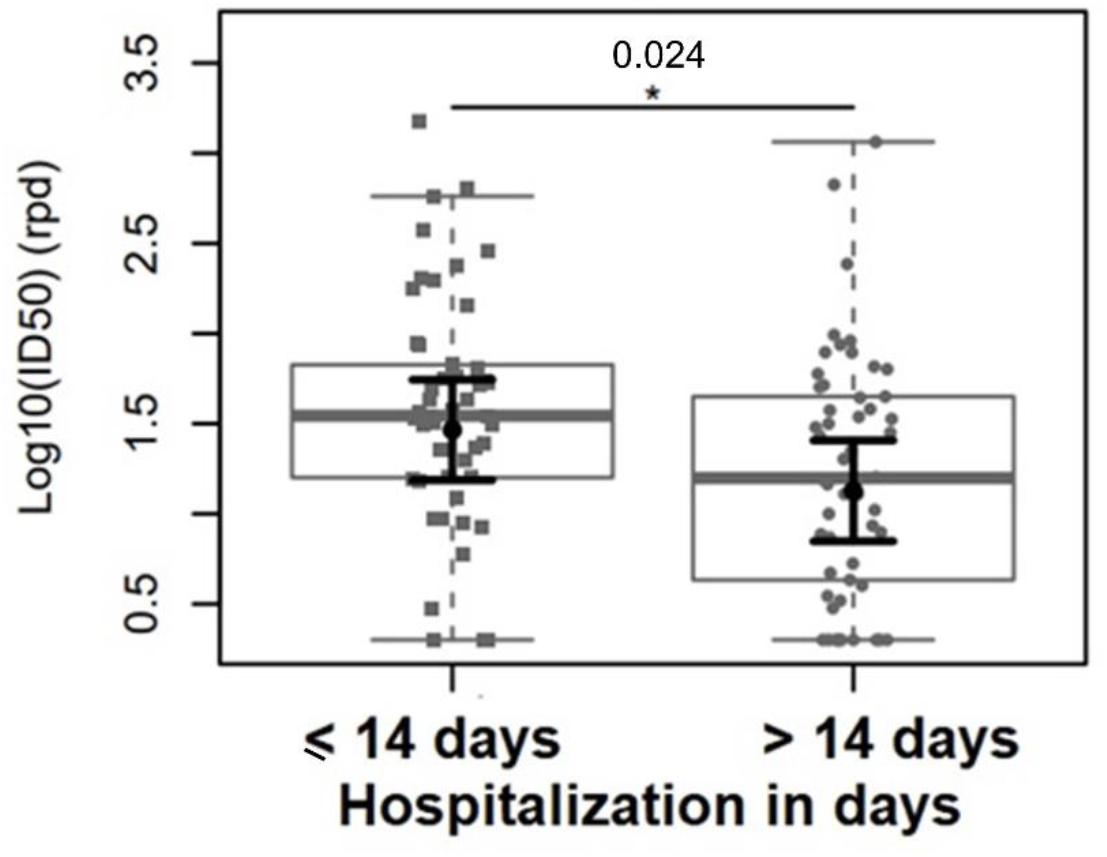
PsV SARS-CoV-2 neutralizing antibody ID_50_reciprocal plasma dilutions and hospitalization stay (in days). The sample distributions of data are represented in box plots and strip plots in gray. In black, the center circle represents the expected mean marginal effect for each group estimated from linear multiple fixed effects models. The fixed effects were adjusted by age, sex, self-declared race, number of comorbidities, and days since the first symptoms of COVID-19 at hospital admission. Black horizontal bars represent the 95% confidence intervals of the expected mean marginal effects by group.

**Figure 3:**
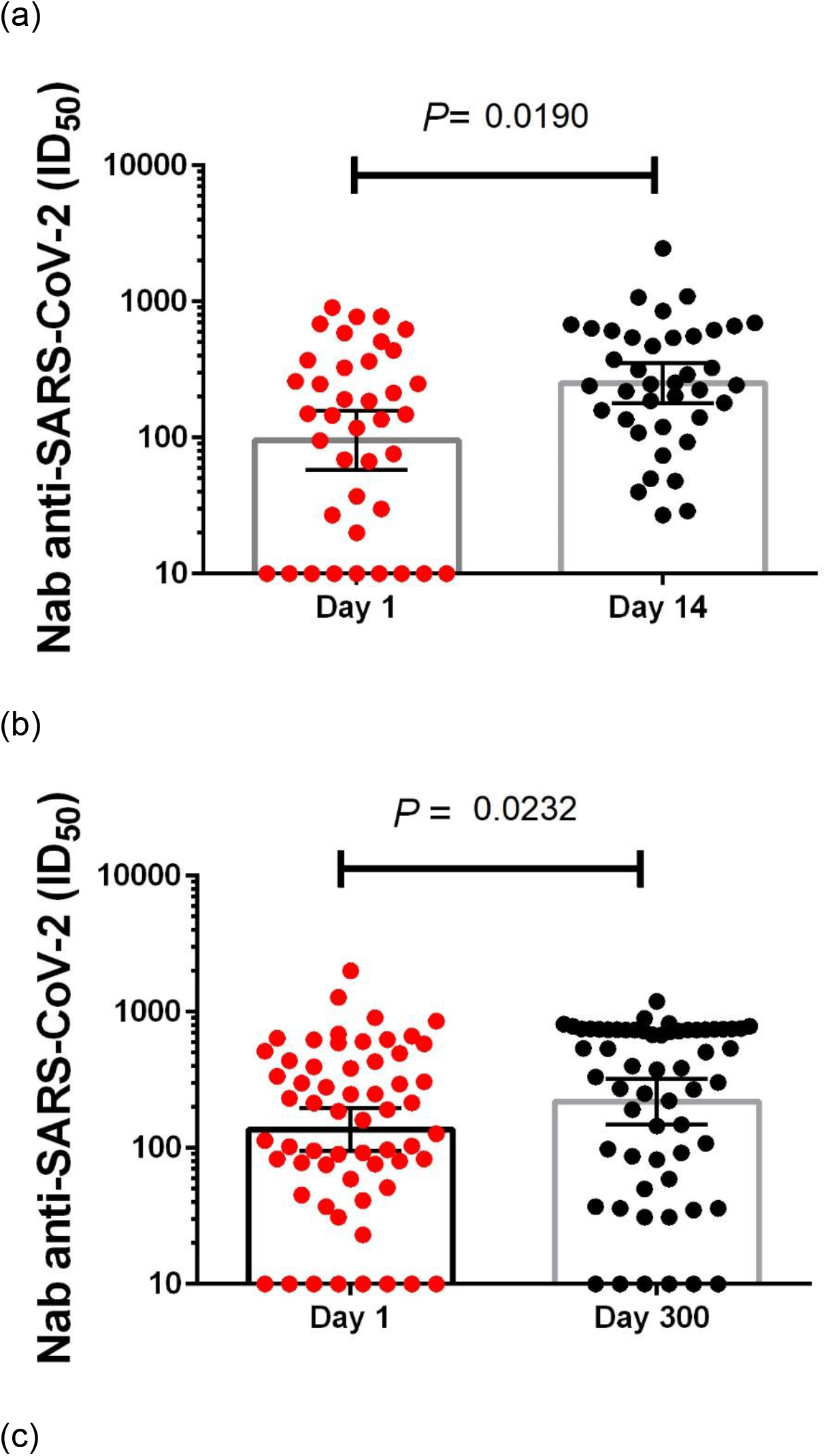
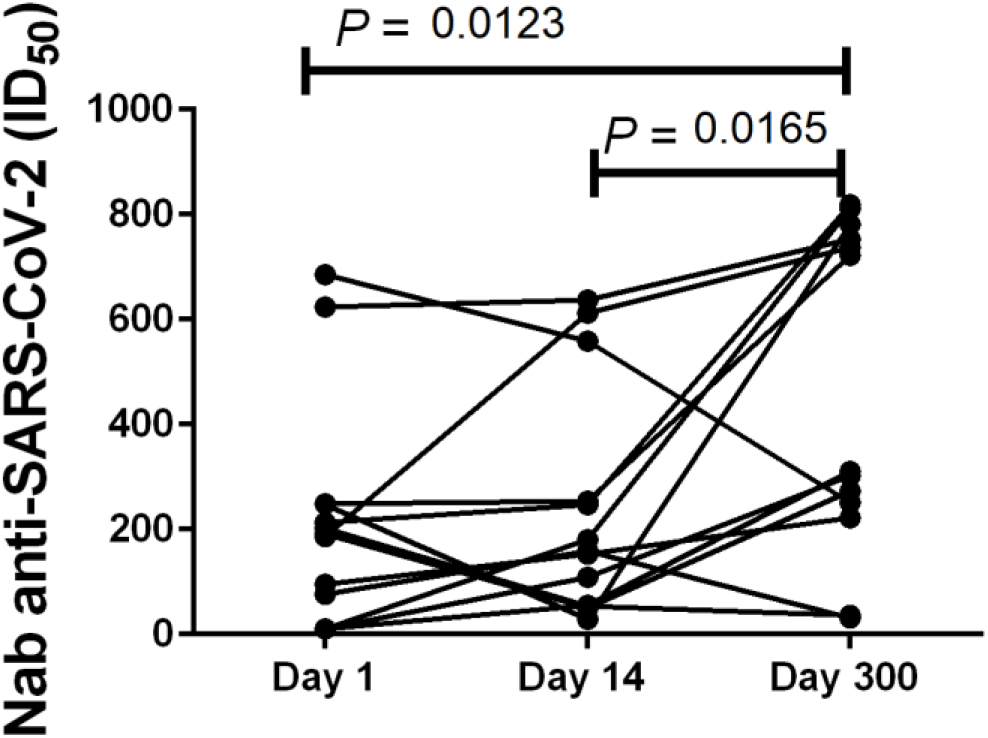
Longitudinal humoral immune response of inpatients with COVID-19. A, Nabs (ID_50_) of COVID-19 patients at admission D1 compared to the Nab (ID_50_) assessment 14 days after hospital discharge. B, Nabs (ID_50_) of COVID-19 patients at admission D1 compared to Nab (ID_50_) assessment 300 days after hospital discharge. C, Changes in Nab titers (ID_50_) for each patient are shown over time. Data from patients for the first sample D1, after 14 days and the last collected sample D300 are shown.

**Figure 4:**
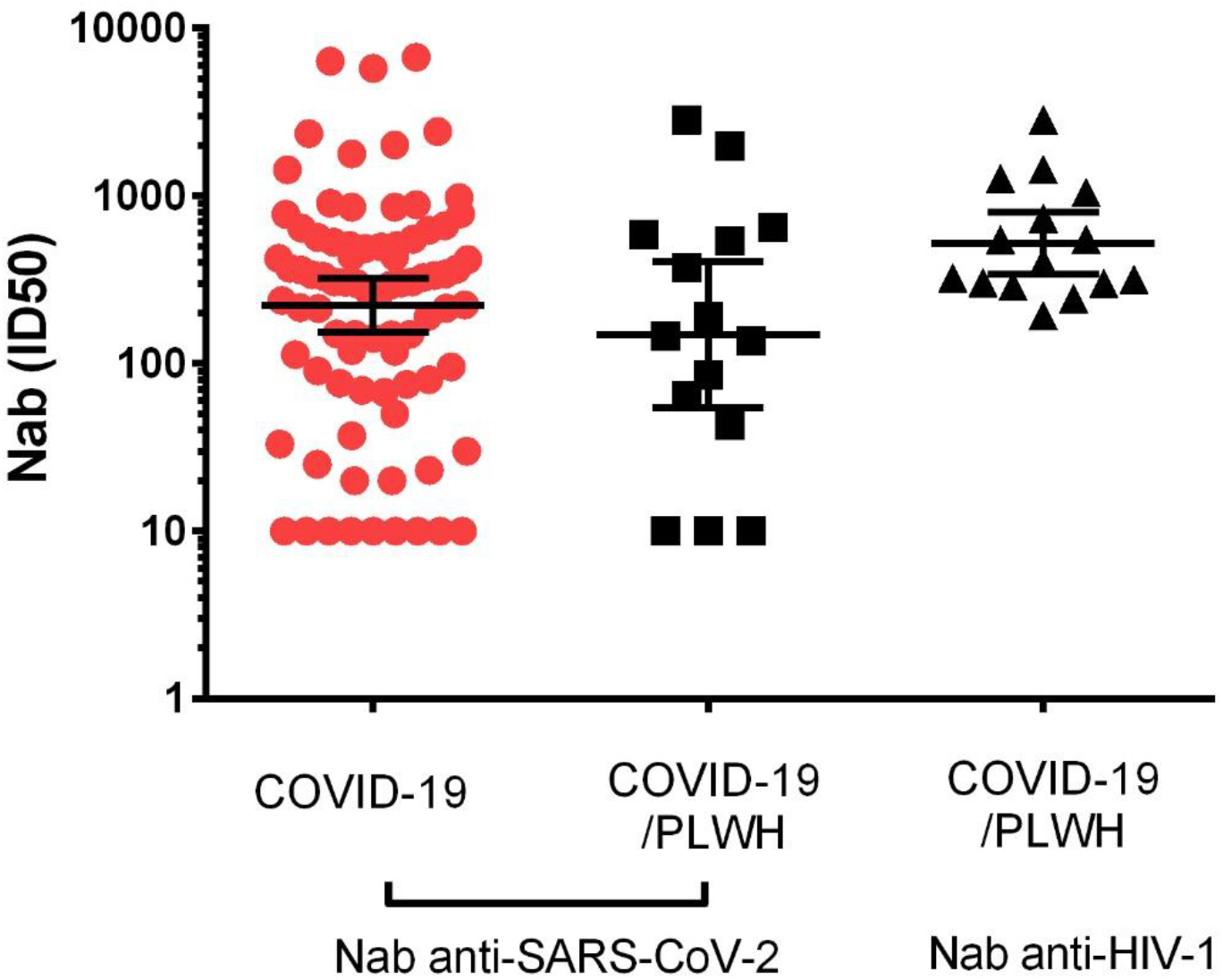
Neutralizing antibody responses from SARS-CoV-2-infected and HIV-coinfected patients. Nabs during the first day of hospitalization of 85 COVID-19 patients not infected with HIV and 15 PLWH coinfected with SARS-CoV-2 (COVID-19/PLWH) are shown. Fifteen patients (COVID-19/PLWH) showed Nab titers against SARS-CoV-2 and HIV-1 pseudovirus. Negative values (titers below 20) were considered 10 for calculating the geometric mean titer (GMT). The horizontal bars indicate the GMT, and the bars indicate 95% confidence intervals.

## 3. Results

### 3.1. Characteristics of the study population

Plasma samples from 103 hospitalized participants were analyzed for Nab response to SARS-CoV-2. Sociodemographic and clinical data were collected for the present study on the first day of hospital admission. Participants were categorized according to the length of hospitalization into short hospitalization (less than 14 days) and long hospitalization (over 14 days) groups to correlate sociodemographic characteristics (Table 1) and clinical data (Table 2). Skin color, age, and sex were not significantly distinct between the short and long hospitalization groups, but 71 (68.9%) participants self-declared that they had brown skin. Most patients were male (57 (55.3%)), and the mean age was 57.27 (IQR=23.51) years (Table 1).

**Table 1.** Sociodemographic characteristics of study participants.

| Characteristics |  | Hospitalization Stay Days |  |  | P-value |
| --- | --- | --- | --- | --- | --- |
|  |  | Overall | 1-14 | >14 |  |
| Total |  | 103 (100%) | 50 (48.6%) | 53 (51.4%) |  |
| Gender | Male | 57 (55.3%) | 28 (56%) | 29 (54.7%) | 1 |
|  | Female | 46 (44.7%) | 22 (44%) | 24 (45.3%) |  |
| Skin Color | Brown | 71 (68.9%) | 30 (60%) | 41 (77.4%) | 0.032 |
|  | White | 14 (13.6%) | 6 (12%) | 8 (15.1%) |  |
|  | Black | 8 (7.8%) | 5 (10%) | 3 (5.7%) |  |
|  | Unk | 10 (9.7%) | 9 (18%) | 1 (1.9%) |  |
| State of Origin | RJ | 101 (98.1%) | 49 (98%) | 52 (98.1%) | 1 |
|  | AM | 2 (1.9%) | 1 (2%) | 1 (1.9%) |  |
| Schooling | High school | 42 (40.8%) | 23 (46%) | 19 (35.8%) | 0.587 |
|  | Elementary School | 20 (19.4%) | 8 (16%) | 12 (22.6%) |  |
|  | University Education | 8 (7.8%) | 4 (8%) | 4 (7.5%) |  |
|  | First grade | 22 (21.4%) | 10 (20%) | 12 (22.6%) |  |
|  | Illiterate | 11 (10.6%) | 5 (10%) | 6 (11.5%) |  |
| Age |  | 57.27<br>(IQR=23.51) | 56.37<br>(IQR=23.59) | 57.57<br>(IQR=25.39) | 0.847 |
| Age Category | 18-39 | 18 (17.4%) | 5 (10%) | 13 (24.5%) | 0.045 |
|  | 40-59 | 34 (33%) | 21 (42%) | 13 (24.5%) |  |
|  | 60-79 | 45 (43.7%) | 23 (46%) | 22 (41.5%) |  |
|  | 80-90 | 6 (5.9%) | 1 (2%) | 5 (9.5%) |  |

Values are presented as number (%) or median (interquartile range).

**Table 2.**
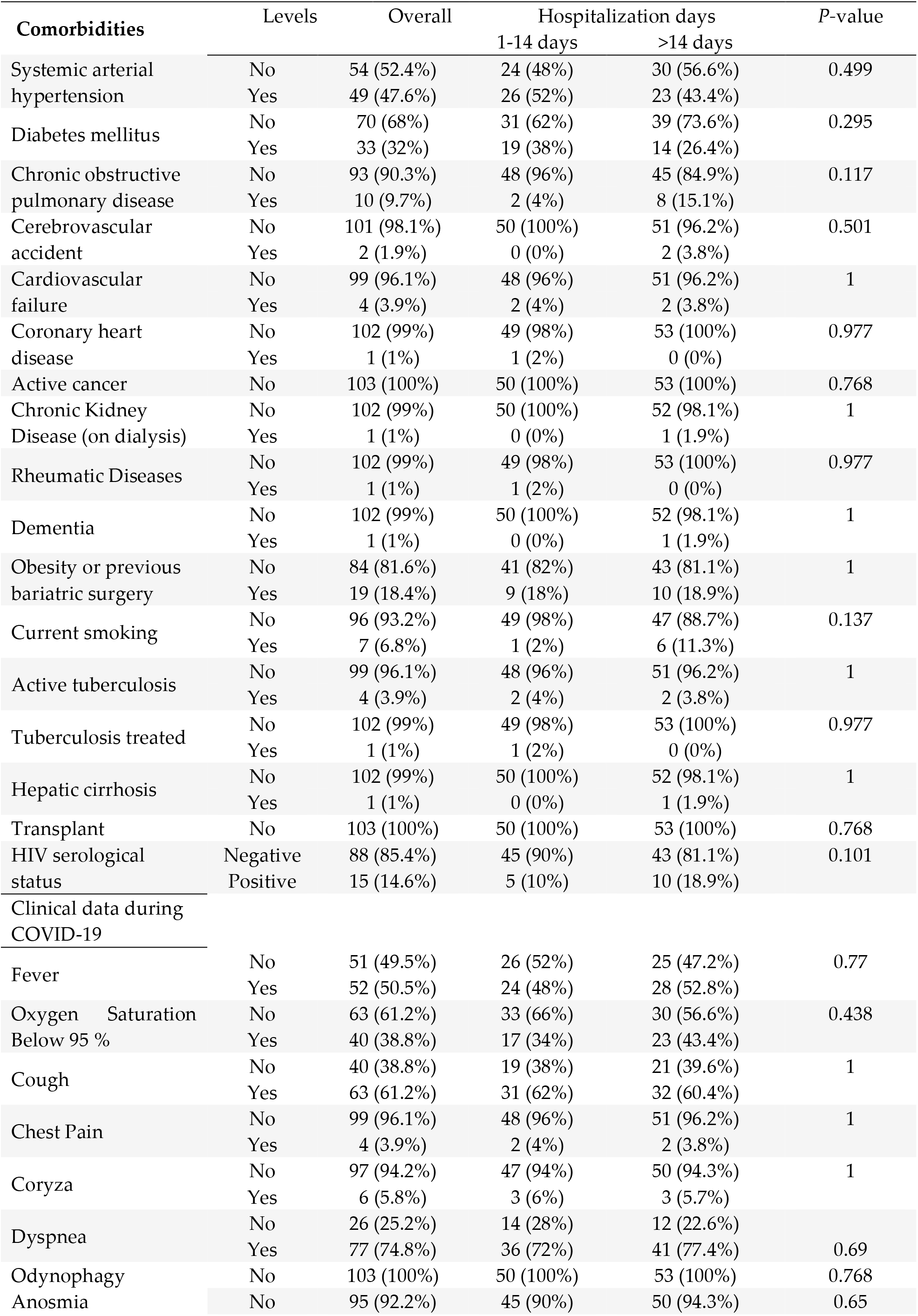
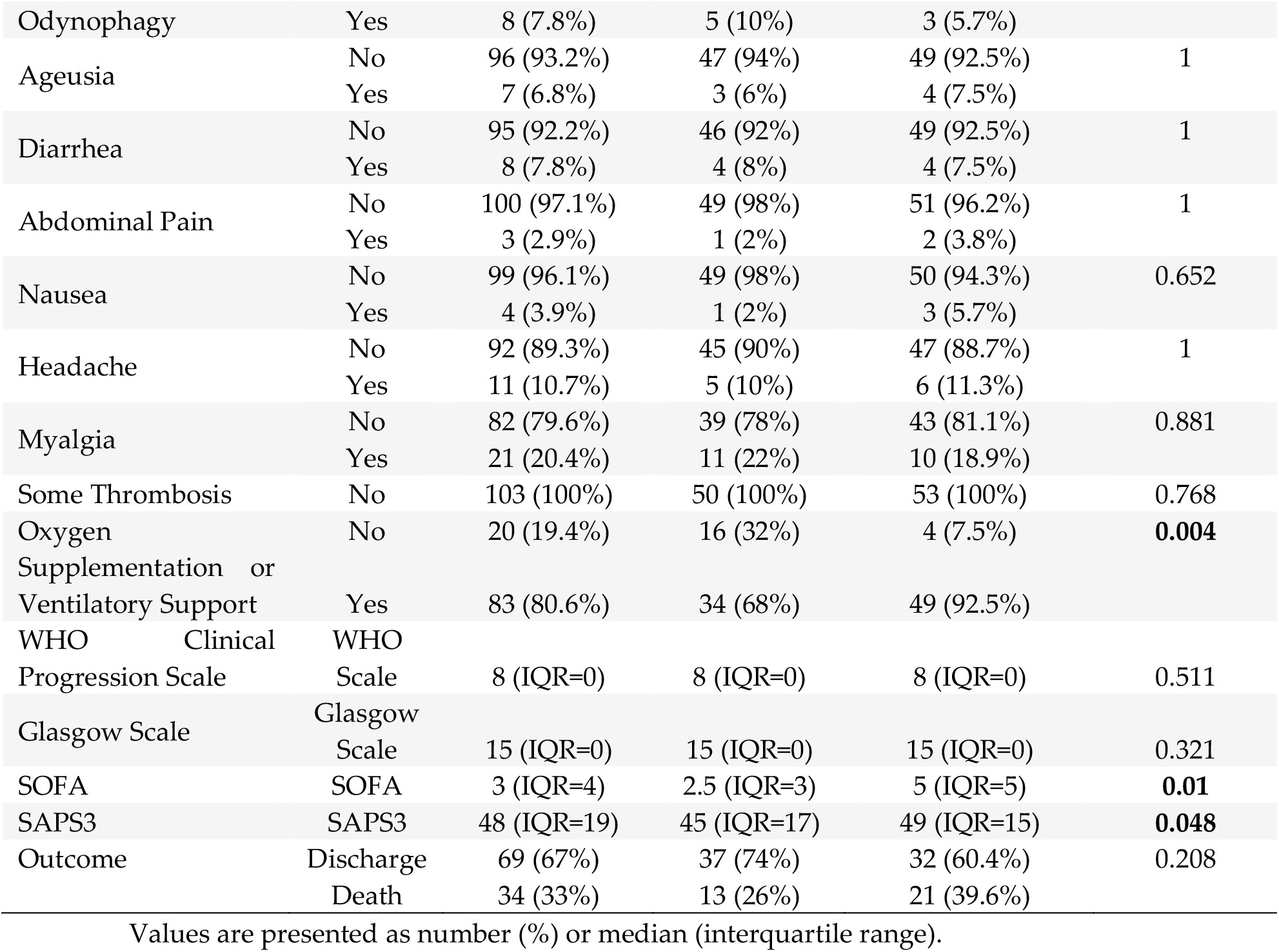
Clinical characteristics and symptoms of study participants.

The most common symptoms observed were fever, cough, chest pain, coryza, dyspnea, anosmia, ageusia, diarrhea, abdominal pain, nausea, headache, myalgia, and oxygen saturation below 95%. The presence of these symptoms varied among the participants, with dyspnea symptoms being the most frequent among the participants (74%), followed by oxygen saturation below 95% (37.5%) and myalgia (21%). We observed that most individuals required oxygen (80.6%) and remained hospitalized for longer periods, *P* <0.01 (Table 2). Major comorbidities included hypertension (49 (47.1%)), diabetes mellitus (33 (31.7%)), and myalgia (22 (21.2%)). Other comorbidities were present in less than 20% of the participants.

Among the individuals included in this study, 15 (14.4%) tested positive for HIV-1 infection (PLWH). Of them, 9 (60%) were male, 14 (93.3%) self-declared having brown skin, and none had obesity, anosmia, loss of taste, nausea, previous stroke, heart failure, or coronary or rheumatic disease. Only 3 (20%) PLWH had systemic arterial hypertension (SAH), which was different from the group without HIV infection, where the majority, 54.1%, had this comorbidity (*P*=0.02). The mean age of patients hospitalized with COVID-19 was 57.27 years. However, among PLWH/people with COVID-19, the mean age was 41.7 years (IQR=17.5), and among those with only COVID-19, the mean age was 59.3 years (IQR=22; *P*<0.01).

### 3.2 Early humoral immune responses among hospitalized patients with severe COVID-19

Humoral anti-SARS-CoV-2 immune responses were assessed at study entry for COVID-19 patients with severe disease. We noted variable levels of Nab anti-SARS-CoV-2 ID_50_in their plasma samples collected on the first day of hospitalization, corresponding to approximately nine days after the onset of the first symptoms. Two patients (1.9%) had very high titers of Nabs (ID_50_) of 1:11,532 and 1:14,962, 17 (16.5%) had high neutralization titers, between 1:500 and 1:10,000 Nab ID_50_, whereas 72 (70%) of the patients developed Nab titers between 1:500 and 1:20. Twelve patients (11.6%) had Nab titers under the detectable limit of the assay (<1:20). Therefore, approximately 88% of COVID-19 patients exhibited robust neutralization of SARS-CoV-2 PNA at hospitalization. Patients who had detectable production of Nabs were hospitalized on average for less time (17 days) than those who did not have detected Nabs (27 days), *P*=0.0363.

As expected, the VSV-G pseudotyped virus control showed the highest infectivity for all samples at a 1:20 dilution.

### 3.3 Association between anti-SARS-CoV-2 neutralizing antibodies and the clinical condition of COVID-19 patients

Interestingly, we found that although the mean age of the patients included in this study, we observed that 18 patients were between 18 and 39 years of age, and of these, 13 (72%) were hospitalized for more than 14 days. The clinical aspects of this youngest group with longer hospitalization were evaluated. Twelve (92.3%) declared themselves as having black skin color, and none of them had comorbidities such as stroke, cardiac insufficiency, coronary artery disease, active cancer, chronic kidney disease or dialysis, rheumatic diseases, dementia, obesity or previous bariatric surgery, hepatic cirrhosis or transplant. However, six (46%) of them were PLWH, and one of them also had active tuberculosis. The Nab geometric mean titer (GMT) ID_50_in this youngest group of patients hospitalized for more than 14 days measured at study entry was 148 (95% CI: 56-387), while the GMT of young people hospitalized for less than 14 days was 360 (95% CI: 83-1,553).

Considering the COVID-19 discharge (n=69) or death (n=34) outcomes in our study group, we observed that the GMT of anti-SARS-CoV-2 Nabs among patients with a death outcome was 140 (95% CI: 74-263), whereas a Nab GMT of 254 (95% CI: 170-380) was observed for those with a discharge outcome. Hospitalization time was longer for patients who died (22; 95% CI: 15-29 days) than for those who were discharged (16; 95% CI: 13-19 days), *P*=0.040. Of the 69 COVID-19 patients who were discharged, 7 (10%) had no Nab response on D1 and were hospitalized for an average time of 20 (95% CI: 10-30) days. Of the 34 patients who died, 5 (14%) had no Nab on D1 and were hospitalized for an average of 35 (95% CI: 13-58) days.

We further explored the clinical manifestations associated with the Nab (ID_50_and ID_90_) levels of the 103 hospitalized patients with COVID-19, and we did not identify any association between the production of Nab titers and the main comorbidities such as arterial hypertension, diabetes, or obesity. However, multivariate analysis showed that anosmia and ageusia (*P*<0.01) were associated with higher Nab titers. All patients with anosmia or ageusia had detectable Nab titers (Figure 1). The Nab GMT of patients who did not have anosmia [32.2 (95% CI: 25-42)] or ageusia [31.7 (95% CI: 24-41)] was lower than that of patients who had anosmia [123 (95% CI: 28-536)] or ageusia [187 (95% CI: 40-869)].

To analyze other relevant clinical factors associated with Nabs, we investigated the hospitalization time in relation to initial Nab production on the first hospital day. We stratified the samples into two groups: 50 (48.6%) patients who stayed for up to 14 days and 53 (51.4%) patients who stayed for more than 14 days. We found significantly lower psV SARS-CoV-2 Nab ID_50_reciprocal plasma dilutions among patients hospitalized for more than 14 days (logFC=0.34; *P*=0.024) than among those with shorter hospitalization stays (Figure 2). The anti-SARS-CoV-2 Nab titer of patients with less than 14 days to hospital discharge [GMT=422 (95% CI: 341-2014)] was significantly higher than that of patients with more than 14 days to hospital discharge [GMT=149 (95% CI: 32-1687)] (*P*<0.05).

To better understand the role of humoral immunity against SARS-CoV-2 infection, we analyzed the dynamics of changes in the Nab response initially at 14 days post-hospitalization (D14) compared to those obtained at hospital admission (D1). As shown in Figure 3A, plasma samples from D14 were available for 39 (73.5%) of the 53 patients who had longer hospital stays (>14 days). There was a significant increase (*P*=0.0190) in Nab titers (ID_50_) between D1 [GMT=95 (95% CI: 57-158)] and D14 [GMT=251 (95% CI: 178-354)].

Patients who were discharged from the hospital were invited to return to the INI/FIOCRUZ after seven months for a new blood collection and clinical follow-up. We performed the Nab assay at this point, hereby called D300, as it was the average day after hospital admission. In this analysis, we compared the Nab titers (ID_50_) of D1 and D300 for the 60 patients who returned for the study, and we observed an increased progression and persistence in the production of Nabs among these SARS-CoV-2-infected and unvaccinated individuals. The GMT for D1 was 136 (95% CI: 94-197), and the GMT for D300 was 219 (95% CI: 149-321) (*P*=0.0232) (Figure 3B).

It was possible to compare D1, D14, and D300 for only 13 patients, and we observed the following values of Nab D1 (ID_50_) GMT=108 (95% CI: 44-266); Nab D14 (ID_50_) GMT=219 (95% CI: 120-400) and Nab D300 (ID_50_) GMT=317 (95% CI: 161-624), (*P*=0.0123 for D1 versus D300; *P*=0.0165 for D14 versus D300) (Figure 3C).

Additionally, we evaluated the geometric mean titers of the anti-SARS-CoV-2 Nab ID_50_of the 15 COVID-19 patients living with HIV-1 (PLWH/COVID-19) included in our study group and compared it with HIV-negative COVID-19 patients. The PLWH/COVID-19 participants [(GMT=148 (95% CI: 54-407)] showed no differences in anti-SARS-CoV-2 Nab production when compared to individuals without HIV-1 infection (GMT=222 (95% CI: 153-321), *P*=0.950 (Figure 4). As a control, we also performed an HIV-1 PNA to determine the anti-HIV Nab titers of the PLWH/COVID-19 participants on antiretroviral therapy. A high anti-HIV-1 antibody GMT of 583.9 (95% CI: 344-990) was detected, demonstrating maintenance of anti-HIV-1 antibody production in individuals with HIV-1 hospitalized for COVID-19.

## 4. Discussion

Age is one of the most important prognostic factors associated with lethality in SARS-CoV-2 infection (11). However, in our study, which included hospitalized patients with moderate to severe COVID-19, age was not shown to be a determining factor for the production or magnitude of anti-SARS-CoV-2 Nabs in adults. In the population evaluated, we observed that 55% of those hospitalized patients were men, the mean age was 57 years, and the mortality rate was 33%. The same was observed in the SIVEP-Gripe study, where 678,235 patients were admitted to Brazilian hospitals and showed a mean age of 59 years and 35% mortality (12). Perazzo et al., 2022, in a prospective multicenter study (RECOVER-SUS) that assessed hospital mortality, analyzed a cohort that included 1,589 participants and found that 54.5% of those admitted were men aged 62 and that 27.0% died during hospitalization (13). The sociodemographic characteristics of the population of our study reflect the data presented by Zeiser et al., 2022 and Perazzo et al., 2022, indicating a consensus on the profile of hospitalized patients with COVID-19 in Brazil. Most men are aged between 50 and 60 years and have approximately 30% mortality.

The most frequent comorbidities observed in our study were hypertension, diabetes, and obesity. Hypertension is more frequent among elderly individuals and subjects affected by other comorbidities (14). In a multivariate analysis after adjustment for age and other cardiovascular risk factors, hypertension did not play an independent role in COVID-19 development (15). Our study observed similar anti-SARS-CoV-2 Nab titers between groups with and without these main COVID-19 comorbidities. However, the survey by Karuna et al., 2021 noted that hypertension was independently associated with low Nab titers (16). According to some researchers, more than half of COVID-19 patients experienced hyperglycemia, and approximately 33% developed diabetic ketoacidosis (17). Marchand et al., 2020 were the first to observe an increased risk of type 1 diabetes development among patients with COVID-19 (18). In our study, the proportion of diabetes patients was 31%, but the presence of diabetes was not related to the ability to produce Nabs, similar to the results observed by Dispinseri et al., 2021 (19).

We observed a correlation between anosmia and ageusia with high Nab titers. The American Academy of Otolaryngology–Head and Neck Surgery (AAO-HNS) released the COVID-19 Anosmia Reporting Tool for Clinicians, which revealed that anosmia was noted in 73% of COVID-19 cases and was the presenting symptom in 26.6% of cases (20). The incidence of anosmia among COVID-19 patients, however, varies in different studies. According to a meta-analysis by Tong et al., 2020, the prevalence of olfactory dysfunction among COVID-19 patients was estimated to be 52% (21). This variability could be due to genetic factors, viral load (cases of viral damage), specificities of the different evaluated populations, or antibody complexes. However, this last concept must be explored since we found higher Nab titers among patients with anosmia. For example, anosmia is one of the first clinical signs of some neurodegenerative diseases, such as Parkinson’s and Alzheimer’s diseases. IgG deposits are found in dopaminergic neurons in these patients, suggesting that neurons may be targets for these immunoglobulins (22). In addition, changes in the blood–brain barrier due to inflammation may facilitate the entry of IgG (23). Jae-Hoon Ko. et al. (2020) evaluated 15 asymptomatic patients who presented only anosmia, and the production of Nabs was observed in 80.0% of patients (24). However, the underlying biological basis of anosmia remains obscure, and our study has some limitations. First, our cohort was limited to hospitalized patients, and the results could be different among people with SARS-CoV-2 infection with mild symptoms. Most hospitalized patients had positive Nab titers (88%) with, on average, nine days of symptoms. Other studies also observed the presence of antibodies in acute infection, with a negativity range in 10-25% of patients (25, 26). Notably, seven out of 12 (58%) recovered patients had Nab titers that were under the detectable level, suggesting that other immune responses may have contributed to the recovery of these patients. Previous studies have suggested that cellular immunity can mitigate severe infection, and T cells display strong cross-reactivity to SARS-CoV-2 (27). Whether these patients with COVID-19 without Nabs have a high risk of rebound or reinfection or a poor response to vaccination should be explored in further studies. Our data revealed that individuals who recovered from COVID-19 experienced relatively robust antibody responses to SARS-CoV-2 PNA in the acute infection phase. However, patients who arrived at the COVID-19 hospital center with a moderate to critical condition and had low titers remained hospitalized for more than 14 days, and 92.5% needed ventilatory support. Therefore, contrary to what we expected, these patients who initially showed a low neutralizing response did not show milder clinical characteristics, such as the asymptomatic individuals with lower antibodies, as seen in other studies (28, 29).

Analysis of the humoral response across multiple cohorts of COVID-19 patients showed that a natural SARS-CoV-2 infection can elicit Nabs in most cases, but accumulating evidence indicates that the magnitude of the response varies greatly among individuals (30, 31). It is still unclear what interferes with the ability to produce potent Nabs. However, it has been observed that plasmablast expansion without somatic hypermutation during the acute phase of COVID-19 plays an important role in the early production of Nabs, which may reflect preexisting B repertoire memory (32).

We observed that after 14 days of hospitalization, Nab GMTs increased in relation to D1, as observed in other cohorts, where the kinetics of Nab induction were reported to be similar for immunoglobulin seroconversion (31). Patients who died of COVID-19 were also able to increase antibody production at D14; however, this delay in the initial immune response may have facilitated the expansion of the viral load, leading to prolonged hospital stays and/or death. Some clinical studies that have adopted the use of convalescent plasma or monoclonal antibodies have observed patient improvement if administered at the beginning of infection (33), and when the serum is administered late (28 days), there was no improvement in the patient (34). The testing of Nabs in the acute phase of COVID-19 can be a great ally for decision-making in clinical trials of convalescent plasma administration as therapy. Another aspect that corroborates the use of Nab identification and not just IgG titration is the presence of antibody-dependent enhancement (ADE), which may be involved in the clinical observation of increased severity of symptoms associated with early high levels of SARS-CoV-2 antibodies in patients (35).

The most extensive study on HIV/COVID-19 coinfection is from a Western Cape, South Africa cohort. PLWH who contracted SARS-CoV-2 died at 2.39 times the rate of patients without HIV with COVID-19. However, HIV was a minor risk factor for severe COVID-19 compared with comorbidities such as cardiopulmonary diseases (36). In our analysis of the production ability of anti-HIV and anti-SARS-CoV-2 antibodies among coinfected patients, we did not observe differences in relation to the mono-infected group (SARS-CoV-2), indicating that HIV-1 is not an aggravating factor for COVID-19 in terms of humoral responsiveness, but we also observed that a low titer of antibodies resulted in a longer hospitalization. Mishra et al., 2021 observed that some HIV-1 Nabs recognize viral glycan shields of SARS-CoV-2, showing cross-reactivity with SARS-CoV-2 (37). How much the polyclonal cross-neutralizing antibody response can contribute to the improvement of the patient has not yet been elucidated; however, in this study, we verified that a potent neutralizing humoral response at the beginning of the infection is associated with a faster recovery.

All protection mechanisms of COVID-19 are not yet clear; however, Nabs are considered a significant correlate of protective immunity and vaccine success, and it has been shown that the rate of SARS-CoV-2 infections among vaccinated individuals is lower among individuals with high Nab levels (38). Extensive efforts to understand the relationship between SARS-CoV-2 and the immune system have highlighted Nab detection assays as essential for screening for potent antibodies for future immunotherapeutic designs. Here, we showed an important relationship between the rapid production of Nabs and shorter hospital stays among patients with moderate and severe COVID-19.

## Author Contributions

Conceptualization, DVA and MGM ; Data curation, MGM, MSBQ, MPR, SWC, BGJG and VGSV; Formal analysis, DVA, MSBQ and MRA; Funding acquisition, DVA, BGJG, VGSV and MGM; Investigation, DVA, LM, CBWG, FHC, MLG, JHP, NBRS, MRA, LEC, KMG, MPR, SWC, BGJG, VGSV and MGM; Methodology, DVA, PAC, TFBF, MGAS, CBWG, FHC, NBRS and ASC; Project administration, DVA and MGM; Resources, LM, CBWG, FHC, MGM; Supervision, DVA, NBRS, CBWG, MGM, VGSV, BGJG; Visualization, PAC, TFBF, MGAS, JHP, ASC, LRG, SWC, LEC, KMG, VGSV and MGM; Writing – original draft, DVA; Writing – review & editing, MGM, CBWG, NDS, MSBQ, ASC, MRA.

## Data Availability

All data produced in the present work are contained in the manuscript

## Funding

The study was supported by the Fundação Carlos Chagas Filho de Amparo à Pesquisa do Estado do Rio de Janeiro (FAPERJ) (Grant numbers SEI-260003/013002/2021) and INOVA FIOCRUZ/Fundação Oswaldo Cruz (Grant numbers SEI-25380.001587/2020-20).

## Institutional Review Board Statement

The study was conducted in accordance with the Declaration of Helsinki and approved by the Ethics Committee of National Institute of Infectology Evandro Chagas (INI)/Oswaldo Cruz Foundation (FIOCRUZ), Rio de Janeiro, Brazil, under the approval number CAAE 32449420.4.1001.5262. All participants or their legal representatives signed an informed consent form prior to enrollment in the study.

## Acknowledgments

The authors are thankful to all patients who agreed to participate in this study, the frontline health care workers at INI/FIOCRUZ Hospital, and the RECOVER study team in Rio de Janeiro. Acknowledgment for publications should read “The following reagent was obtained through the NIH HIV Reagent Program, Division of AIDS, NIAID, NIH: TZM-bl Cells, ARP-8129, contributed by Dr. John C. Kappes, Dr. Xiaoyun Wu and Tranzyme Inc.”

## Conflicts of Interest

The authors declare that the research was conducted in the absence of any commercial or financial relationships that could be construed as a potential conflict of interest. The funders had no role in the design of the study; in the collection, analyses, or interpretation of data; in the writing of the manuscript; or in the decision to publish the results.

## Notes

### Competing Interest Statement

The authors have declared no competing interest.

### Funding Statement

The study was supported by FAPERJ/BR (Grant numbers SEI-260003/013002/2021) and INOVA FIOCRUZ/BR (Grant numbers SEI-25380.001587/2020-20).

### Author Declarations

The study was conducted in accordance with the Declaration of Helsinki and approved by the Ethics Committee of the National Institute of Infectology Evandro Chagas (INI)/ Oswaldo Cruz Foundation (FIOCRUZ), Rio de Janeiro, Brazil, under the approval number CAAE 32449420.4.1001.5262. All participants or their legal representatives signed an informed consent form prior to enrollment in the study.

